# Therapeutic effects of dipyridamole on COVID-19 patients with coagulation dysfunction

**DOI:** 10.1101/2020.02.27.20027557

**Authors:** Xiaoyan Liu, Zhe Li, Shuai Liu, Zhanghua Chen, Jing Sun, Zhiyao Zhao, Yi-you Huang, Qingling Zhang, Jun Wang, Yinyi Shi, Yanhui Xu, Huifang Xian, Rongli Fang, Fan Bai, Changxing Ou, Bei Xiong, Andrew M Lew, Jun Cui, Hui Huang, Jincun Zhao, Xuechuan Hong, Yuxia Zhang, Fuling Zhou, Hai-Bin Luo

## Abstract

The human coronavirus HCoV-19 infection can cause acute respiratory distress syndrome (ARDS), hypercoagulability, hypertension, extrapulmonary multiorgan dysfunction. Effective antiviral and anti-coagulation agents with safe clinical profiles are urgently needed to improve the overall prognosis. We screened an FDA approved drug library and found that an anticoagulant agent dipyridamole (DIP) suppressed HCoV-19 replication at an EC50 of 100 nM *in vitro*. It also elicited potent type I interferon responses and ameliorated lung pathology in a viral pneumonia model. In analysis of twelve HCoV-19 infected patients with prophylactic anti-coagulation therapy, we found that DIP supplementation was associated with significantly increased platelet and lymphocyte counts and decreased D-dimer levels in comparison to control patients. Two weeks after initiation of DIP treatment, 3 of the 6 severe cases (60%) and all 4 of the mild cases (100%) were discharged from the hospital. One critically ill patient with extremely high levels of D-dimer and lymphopenia at the time of receiving DIP passed away. All other patients were in clinical remission. In summary, HCoV-19 infected patients could potentially benefit from DIP adjunctive therapy by reducing viral replication, suppressing hypercoagulability and enhancing immune recovery. Larger scale clinical trials of DIP are needed to validate these therapeutic effects.

## Introduction

The novel coronavirus HCoV-19 (also known as SARS-CoV-2) outbreak had emerged from Wuhan, Hubei Province, China in December 2019.^1,2^ As of February 24, there had been 77,787 confirmed COVID-19 cases including 2,666 deaths in China. Over 2,500 infections have also been reported in 28 other countries, such as Japan, South Korea, Italy, Singapore, Iran, Thailand, and U.S.A; this rapid spread threatens to become a pandemic. To date, no agents have been reported to be effective against this infection. Identification of readily available drugs for repurpose in COVID-19 therapy avail a relatively rapid way to clinical treatment.^3^

HCoV-19, together with SARS-CoV and MERS-CoV, belong to the *Beta-coronovirus* genus, which are enveloped, positive-stranded RNA viruses with approximately 30,000 nucleotides.^4,5^ Angiotensin I converting enzyme 2 (ACE2) is the receptor that engages the Spike surface glycoprotein of SARS-CoV and HCoV-19.^6,7^ ACE2 is expressed in many organs, including the lung, heart, kidney, and intestine. Notably, in experimental models of SARS-CoV infection, Spike protein engagement decreases ACE2 expression and activates the renin-angiotensin system (RAS).^6^ RAS activation promotes platelet adhesion and aggregation and increases the risk for pulmonary embolism, hypertension and fibrosis.^8-11^ It also accelerates cardiac and kidney injury by increasing local angiotensin II concentrations.^12-14^ Apart from affecting the classic RAS pathway, ACE2 deficiency in the intestine is associated with malnutrition and colonic inflammation.^15^

Infection from SARS-CoV can result in severe lymphopenia, prolonged coagulation profiles, lethal acute respiratory distress syndrome (ARDS), watery diarrhea, cardiac disease and sudden death.^9,16-18^ Many of these features have also been reported for COVID-19: prolonged coagulation profiles, elevated D-dimer levels, severe lymphopenia, ARDS, hypertension, and acute heart injury in ICU-admitted patients.^2,19^ Given that angiotensin II levels were highly elevated in the HCoV-19 infected patients, ^20^ RAS was likely activated in HCoV-19 infected patients, Thus, prophylactic anti-coagulation therapy should be considered for mollifying the multi-organ damage during severe COVID-19.

After viral entry to the host cells, the coronavirus messenger RNA is first translated to yield the polyproteins, which are subsequently cleaved by two viral proteinases, 3C-like protease (3CLP, aka nsp5 or Mpro) and papain-like protease (PLP, or nsp3), to yield non-structural proteins essential for viral replication.^21^ Inhibitors that suppress the activity of these proteases may inhibit viral replication and offer a revenue for the HCoV-19 therapy.

Dipyridamole (DIP) is an antiplatelet agent and acts as a phosphodiesterase (PDE) inhibitor that increases intracellular cAMP/cGMP.^22^ Apart from the well-known antiplatelet function, DIP may provide additional therapeutic benefits to COVID-19 patients. First, published studies,^23-28^ including clinical trials conducted in China,^29-31^ have demonstrated that DIP has a broad spectrum antiviral activity, particularly efficacious against the positive-stranded RNA viruses.^24^ Second, it suppresses inflammation and promotes mucosal healing. ^32^ Third, as a pan-PDE inhibitor, DIP may prevent acute injury and progressive fibrosis of the lung, heart, liver, and kidney.^33^ Here we provide three levels of evidence advocating DIP as a therapy. *In silico and in vitro*, we demonstrated that DIP possessed anti-HCoV-19 effects. In a VSV-induced pneumonia model, we also confirmed that DIP elicited potent antiviral immunity and significantly improved the survival rate. In a small clinical cohort, we found that DIP adjunctive therapy led to increased circulating lymphocyte and platelet counts and lowered D-dimer levels, and markedly improved clinical outcomes.

## Methods

### Free energy perturbation and surface plasmon resonance (SPR) assay

We virtually screened an FDA-approved drug database using the HCoV-19 protease Mpro as a target and validated the binding affinity by the SPR assay. DIP (PubChem CID: 3108, Figure S1) was identified as a lead drug. In order to obtain the binding pattern and calculate the binding free energy between DIP and Mpro, DIP was firstly docked onto Mpro by using Glide, and the optimal binding pose was further assessed by absolute binding free energy calculation with free energy perturbation. ^34^ The calculations were carried out in Gromacs 2019, and the thermodynamic cycle and procedure was similar to that used by Matteo et al.^35^ In the calculation, the ligand electrostatic and van der Waals interactions were decoupled using a linear alchemical pathway with Δλ = 0.10 for the van der Waals and Δλ = 0.20 for electrostatic interactions. To add the restraints, 12 λ values were used. Therefore, a total of 28 windows for complex and 16 windows for ligand systems were employed, and 4 ns simulations were performed for each window. The relative position restraints consist of one distance, two angles, and three dihedrals harmonic potentials with a force constant of 10 kcal/mol/Å^2^ [rad^2^]. The distance and angles for the restraints were determined by the values of the last 2 ns of the 4 ns preliminary MD simulations. The sampled *ΔU* in the simulations were fitted by Gaussian algorithms and the free energy estimates were obtained by using the Bennet acceptance ratio (BAR) method.^36^

The pGEX4T1-Mpro plasmid was constructed (Atagenix, Wuhan) and transfected the E. coli strain BL21 (Codonplus, Stratagene). GST-tagged protein was purified by GST-glutathione affinity chromatography and cleaved with FXa. The purity of recombinant protein was greater than 95% as judged by SDS–PAGE. The binding of DIP to Mpro was measured by the Biacore 8K system (GE healthcare) at 25 °C. Mpro was immobilized on a CM5 chip surface via covalent linkage to Mpro N-terminus. First, the CM5 chip was activated using 1:4 *N*-hydroxysuccinimide (NHS)/1-ethyl-3-(3-dimethylaminopropyl) carbodiimide (EDC) at the flow rate of 10 *µ*L/min for 7 min. Then, Mpro (90 *µ*g/mL) in 10 mM acetate buffer (pH 4.5) was passed over separate flow cells at 10 *µ*L/min for 3 min (1800 response units), which followed by a blocking step u sing ethanolamine (1 M, pH 8.5) at 10 *µ*L/min for 7 min. Binding studies were performed by passing 5-80 *µ*M of DIP over the immobilized Mpro at the flow rate of 30 *µ*L/min and the contact time was set to be 200 s. A sample volume of 120 *µ*L DIP in running buffer was injected into the flow cell and the bound ligand was washed by running buffer which contained 10 mM phosphate, 137 mM NaCl, 2.7 mM KCl, pH 7.4, and 0.5% DMSO.

### Fluorescent focus assay

Vero E6 cells were seeded in 96-well plates and treated with different dosages of dipyridamole or chloroquine for an hour before infected with HCoV-19. After 24 hours of incubation, cells were fixed and permeabilized with Cytofix/Cytoperm (BD), incubated with a rabbit anti-HCoV-19 nucleocapsid protein polyclonal antibody (Sino Biological), followed by a HRP-labelled goat anti-rabbit secondary antibody (Jackson). The foci were visualized by TRUEBlue reagent. Foci in infected cells were counted with a ELISPOT reader. Viral titers were calculated as foci forming units (FFU) per ml.

### RNA extraction and quantitative real-time PCR

Total cellular RNA was isolated by TRIzol reagent (Invitrogen) and reverse transcription was performed using a reverse transcription kit (Vazyme). We performed real-time PCR with SYBR Green qPCR Mix (GenStar). Data were normalized to the *RPL13A* gene and relative abundance of transcripts was calculated by the *Ct* models. Following primers were used for real-time PCR:

*IFN-β* forward primer: 5’CAGCAATTTTCAGTGTCAGAAGC3’;

*IFN-β* reverse primer: 5’TCATCCTGTCCTTGAGGCAGT3’;

*ISG15* forward primer: 5’ TCCTGGTGAGGAATAACAAGGG3’;

*ISG15* reverse primer: 5’ GTCAGCCAGAACAGGTCGTC3’;

*IFIT1* forward primer: 5’ TCAGGTCAAGGATAGTCTGGAG3’;

*IFIT1* reverse primer: 5’ AGGTTGTGTATTCCCACACTGTA3’.

*RPL13A* forward primer: 5’GCCATCGTGGCTAAACAGGTA3’

*RPL13A* reverse primer: 5’GTTGGTGTTCATCCGCTTGC3’

### Immunoblotting

Whole-cell extracts were eluted with 5×SDS Loading Buffer and then resolved by SDS-PAGE. The resolved protein bands were electro-transferred to polyvinylidene fluoride membranes for further antibody incubation and detection. The antibodies against TBK1 [TBK1/NAK (D1B4) Rabbit mAb #3504, Phospho-TBK1/NAK (Ser172) (D52C2) XP^®^ Rabbit mAb #5483] and IRF3 [IRF-3 (D83B9) Rabbit mAb #4302, Phospho-IRF-3 (Ser386) (E7J8G) XP® Rabbit mAb #37829] were purchased from Cell Signaling Technology and anti-β-actin (A1978) were purchased from Sigma.

### VSV-induced viral pneumonia model

C57/BL-6J mice were obtained from the Guangdong Medical Laboratory Animal Center. Six-week old female mice were intravenous injected with VSV (10^8^ PFU/g) and followed with DIP (35 mg/kg; intraperitoneal injection, i.p; twice/day) treatment for 5 days. Survival curve was measured. Mice were sacrificed at Day 4 for histology examination by hematoxylin-eosin (H&E) staining. The study procedure was approved by Animal Ethics Committee of Guangzhou Medical University.

### Patients and study variables

As of February 8, 2020, 124 confirmed COVID-19 cases had been identified from Zhongnan Hospital of Wuhan University (**Table S1**). All patients met the diagnostic criteria of “Diagnosis and Treatment Scheme of Novel Coronavirus–Infected Pneumonia (trial 6th)” formulated by the General Office of National Health Committee.^37^ A retrospective review of the medical records of these patients was conducted to retrieve coagulation indexes and platelet parameters, including prothrombin time (PT), activated partial thromboplastin time (APTT), plasma fibrinogen (FIB), D-dimer, platelet (PLT) count, and mean platelet volume (MPV). Systemic inflammation was assessed according to the C-reactive protein (CRP), procalcitonin (PCT), and interleukin 6 (IL-6) levels.^12^

### Study design

Patients were enrolled from the isolation ward in Dawu County People’s Hospital, a medical treatment alliance of Zhongnan Hospital, Hubei province in China, from Jan 23, 2020 to Feb 22, 2020 to determine the effect of DIP to improve the coagulation profile. The condition of the patients was monitored daily by attending physicians. Routine laboratory test of the coagulation variables, blood indexes, liver and kidney function indices, and chest CT were carried out before, during, and after the treatment. Clinical symptoms and laboratory data were independently validated by two independent investigators for assurance of data accuracy. The diagnosis of severe case was made if patients met any of the following criteria: (1) respiratory rate ≥ 30 breaths/min; (2) SpO2 ≤ 93% while breathing room air; (3) PaO2/FiO2 ≤ 300 mmHg. A critically ill case was diagnosed if any of the following criteria was met: (1) respiratory failure which requiring mechanical ventilation; (2) shock; (3) combined with other organ failure, need to be admitted to ICU. The Ethics Committee from Dawu County People’s Hospital approved the study. All patients signed informed consents.

### Treatment procedures

Anticoagulant therapy was provided via oral DIP tablets. The daily treatment protocol comprised of 150 mg in three separate doses for seven consecutive days. All patients were monitored daily for possible adverse events. All patients received antiviral (ribavirin, 0.5g, Q12h), corticoid (methylprednisolone sodium succinate, 40 mg, qd), oxygen therapy, and nutritional support as necessary.

### Statistical analysis

Statistical analyses and graphics production were performed using R v3.5.3 (Foundation for Statistical Computing) and GraphPad Prism 8. Categorical variables were described as frequencies or percentages, and continuous variables were shown as mean with standard deviation/error or median with interquartile range. Comparison for two independent groups was conducted using Student’s *t* test (for normally distributed data) or Mann-Whitney test (for non-normally distributed data). *P* < 0.05 was considered statistically significant. Detailed descriptions of data comparison and statistical tests were specified in the figure legends.

## Results

### DIP suppresses HCoV-19 replication in Vero E6 cells

We virtually screened an FDA approved drug library and found that DIP bound to the HCoV-19 protease Mpro (**Figure S1)**. Hydrophobic and hydrogen bond (H-bond) interactions are the main driving forces for the binding between DIP and Mpro. According to the free energy perturbation calculation, the binding free energy of DG_pred_ was -6.34 kcal/mol. An SPR assay was carried out to validate the *in silico* result. This has revealed that DIP bound to Mpro with an experimentally confirmed affinity of 34 µM (K_*D,eq*_) (**Figure 1A-B**).

**Figure 1.**
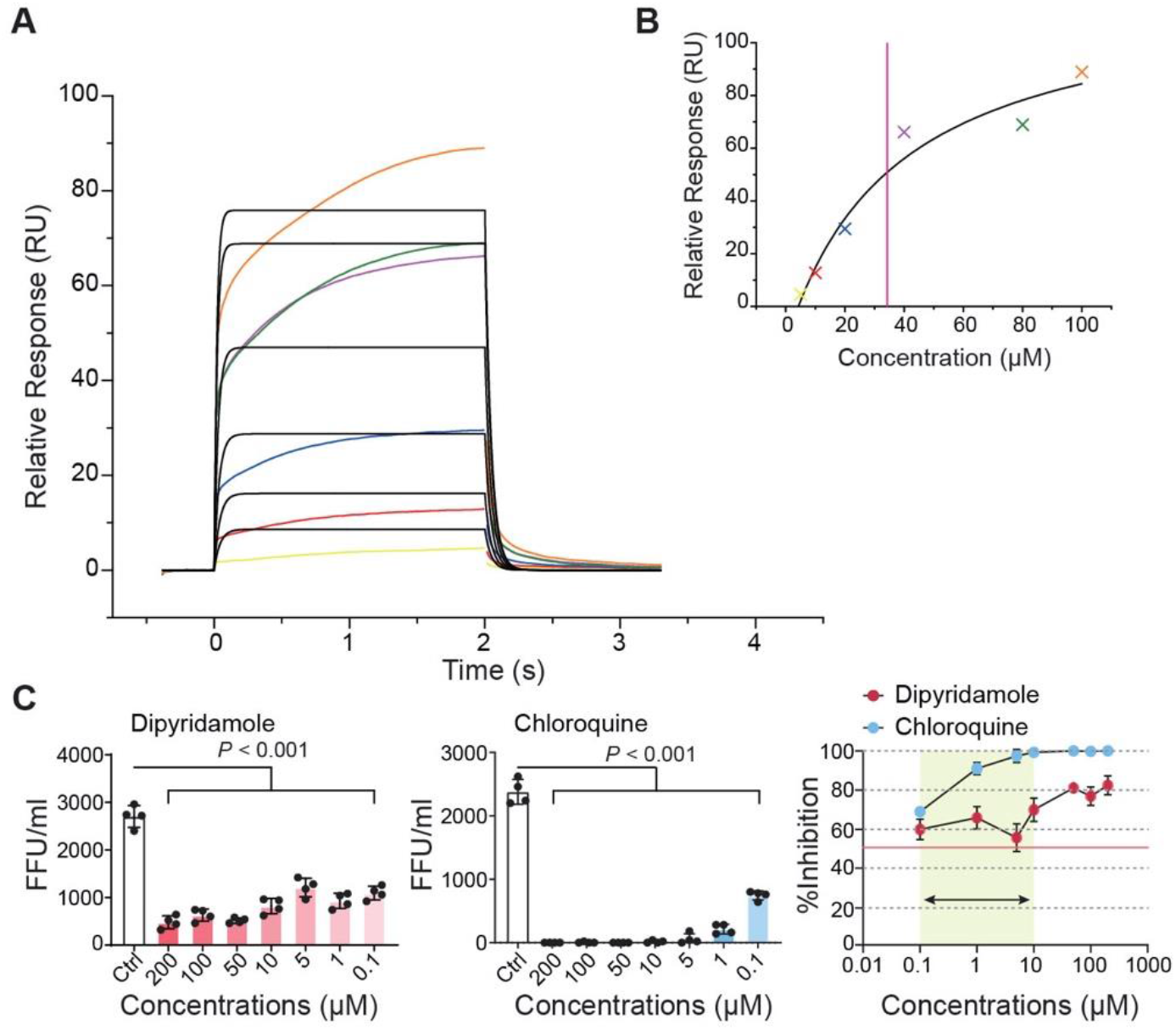
Dipyridamole suppresses Mpro activity by the SPR assay. Experimental SPR sensorgram from Biacore 8K between dipyridamole (DIP) and Mpro, color points overlaid with a 1:1 Langmuir binding model kinetic fit (black). (A) Relative response from injection of DIP at 5, 10, 20, 40, 80, and 100 μM. Global analysis of the shape of the response curves yielded the parameters of on-rate (*K*_*on*_ = 3.20E+3), off-rate (*K*_*off*_ = 2.19E-1) and the ratio of *K*_*off*_ divided by *K*_*on*_ (*K*_*D*_ = 68 µM).(B)Equilibrium binding responses plotted versus DIP concentration and fitted to a simple binding isotherm to yield an affinity of 34 µM (K_*D,eq*_). (C) Suppressive effects of DIP and chloroquine on HCoV-19 replication. *P* values were calculated by Student’s *t* test.

To demonstrate directly that DIP suppresses HCoV-19 replication, we measured the viral titers using a susceptible cell line, the Vero E6 cells. Chloroquine was used as a positive control. ^38,39^ Remarkably, at 0.1 mM DIP suppressed more than 50% of HCoV-19 replication (**Figure 1C**). Human DIP administration can achieve a 3 µM serum concentration.^40^ Thus, therapeutic dosages of DIP may effectively suppress HCoV-19 replication in the infected patients. Notably, however, the dose-dependent suppressive effect of dipyridamole was less apparent in comparison to Chloroquine, which suggested that DIP may employ a different mode of suppression.

### DIP promotes type I interferon (IFN) responses and protect mice from viral pneumonia

Published studies have shown that DIP possessed broad spectrum activity to a wide range of viruses, indicating that it may elicit antiviral immunity from the host cells. We examined these using two single-stranded RNA viruses (Sendai virus, SeV and Vesicular stomatitis virus, VSV), as well as intracellular poly (I:C) that stimulates the RIG-I and MDA5 mediated IFN immunity, in the human lung epithelial cell line A549 cells.^41,42^ Our results demonstrated that DIP (5 μM) treatment significantly increased *IFNB, IFIT1*, and *ISG15* messenger RNA expression and promoted IFN-b secretion (**Figure 2A**) by increasing TBK1 and IRF3 phosphorylation (**Figure 2B**).

**Figure 2.**
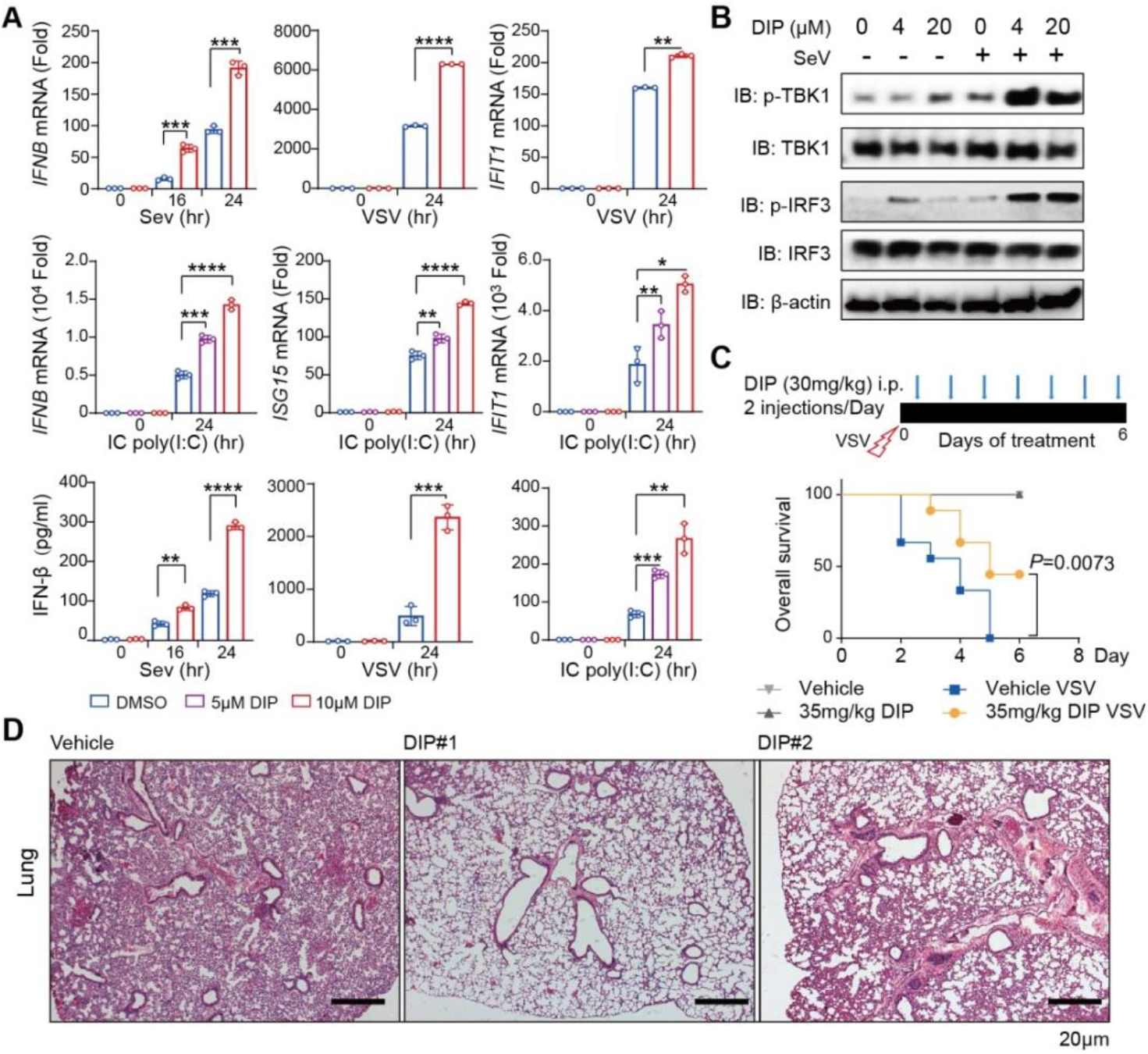
Dipyridamole (DIP) potentiate antiviral immunity to single-stranded RNA viruses. (A) Top, detection of *IFNB* and *IFIT1* mRNA expression in A549 cells treated with or without 5 mM DIP followed by SeV or VSV infection. Middle, real-time PCR detection of *IFNB, ISG15* and *IFIT1* mRNA expressions by A549 cells treated with different doses of DIP (0, 5 and 10 mM), followed by intracellular poly (I:C) treatment. Bottom, ELISA detection of IFN-a production by A549 cells. (B) Immunoblot detection of total and phosphorylated (p-) TBK1 and IRF3 in 293T cells with different doses of DIP (0, 4, 20 mM), followed with or without SeV infection. (C) Schematic of the VSV-infection mice model and the survival curve. (D) Histopathology of lungs from mice sacrificed at Day4. Scale bars, 20 mm. Data in A are presented as the means ± SE of at least three biological experiments. *P* values were calculated by Student’s *t* test, \**P* < 0.05, ** *P* < 0.01, *** *P* < 0.001 and **** *P* < 0.0001. Data in F are presented with 3 to 9 mice per group, *P* value was calculated by Log-rank (Mantel-Cox) test (conservative).

To validate the antiviral effects of DIP *in vivo*, we established a VSV-induced pneumonia model. DIP administration significantly prolonged the survival rate of VSV-infected mice (**Figure 2C**), and substantially alleviated pulmonary pathology (**Figure 2D**). Thus, therapeutic doses of DIP can elicit potent and broad-spectrum antiviral immunity to single-stranded RNA viruses.

### A remarkable improvement of coagulation profile in COVID-19 patients receiving DIP adjunctive therapy

To assess the coagulation profile, we first retrospectively analyzed a randomly collected cohort of 124 COVID-19 patients (**Table S1**). Decreased platelet counts were observed in 25 (20.2%) patients, prolonged prothrombin time (PT) was observed in 77 (62.1%) patients, fibrinogen (FIB) levels were increased in 27 (21.8%) patients, and D-dimer levels were increased in 26 (21.0%) patients. This suggest that hypercoagulability is common in COVID-19 patients.

To evaluate DIP as a therapy to reduce the risk of hypercoagulability, 22 additional patients including 10 control patients and 12 patients who received DIP were recruited. Baseline characteristics of the two groups were similar (**Table 1**). The average ages of the patients were 53 and 58 years in the DIP group and the control group, respectively. All patients manifested cough and most of them felt shortness of breath, and 60.0% patients had nausea and vomiting. Chest CT scan revealed bilateral pneumonia in all patients. Comorbidities, including diabetes mellitus, cardiovascular and cerebrovascular diseases, were found in 4 and 5 of the patients from the DIP and control group, respectively. All patients received ribavirin, glucocorticoids, and oxygen therapy, but none received antifungal treatment. Other treatment included mechanical ventilation (16.7% vs 10.0%), antibiotics (33.3% vs 30.0%), and intravenous immunoglobulin (16.7% vs 20.0%) in the DIP group and the control group, respectively.

**Table 1.** Baseline characteristics of enrolled patients.

| **Table 1. Baseline characteristics of enrolled patients** |  |  |
| --- | --- | --- |
| **Variable** | **Dipyridamole group**<br>**(n=12)** | **Control group**<br>**(n=10)** |
| **General Characteristic** |  |  |
| Age (yr) — mean±sd (range) | 53±10 (32-69) | 58±15 (23-74) |
| Gender— male no. / female no. | 8 / 4 | 7 / 3 |
| **Clinical variables** |  |  |
| Cough— no. (%) | 12 (100.0%) | 10(100.0%) |
| Shortness of breath — no. (%) | 9 (75.0%) | 7(70.0%) |
| Nausea and vomiting— no. (%) | 7 (58.0%) | 6(60.0%) |
| Systolic blood pressure (mmHg) | 125±13(120-155) | 128±11(110-150) |
| — mean±sd (range) | /77±11(57-98) | /79±5(70-90) |
| Heart rate (beats/min) — mean±sd (range) | 87±7(75-100) | 83±12(59-101) |
| Partial pressure of oxygen (mmHg) | 86±12 (58-93) | 92±9 (76-96) |
| — mean±sd (range) |  |  |
| **Laboratory values** |  |  |
| WBC (10<sup>9</sup>/L) — mean±sd (range) | 6.84±3.07 (3.65-12.74) | 5.19±0.72 (3.96-5.92) |
| Lymphocyte (10<sup>9</sup>/L) — mean±sd (range) | 1.11±0.61 (0.29-2.28) | 1.00±0.49 (0.17-1.85) |
| D-dimer (mg/L) — mean±sd (range) | 2.35±2.91 (0.19-6.84) | 1.54±2.86 (0.17-8.43) |
| Increase in level of D-dimer — no. (%) | 4 (33.3%) | 2 (20.0%) |
| **Respiratory pathogens** |  |  |
| The nucleic acid of COVID-19 — no. (%) | 12(100.0%) | 10(100.0%) |
| Unilateral pneumonia — no. (%) | 0 | 0 |
| Bilateral pneumonia — no. (%) | 12(100.0%) | 10(100.0%) |
| **Comorbidities** |  |  |
| Diabetes mellitus — no. (%) | 2(16.7%) | 1(10.0%) |
| Cardiovascular disease — no. (%) | 1(8.3%) | 3(50.0%) |
| Cerebrovascular disease — no. (%) | 1(8.3%) | 1(10.0%) |
| **Treatment** |  |  |
| Oxygen therapy — no. (%) | 12(100.0%) | 10(100.0%) |
| Mechanical ventilation — no. (%) | 2(16.7%) | 1(10.0%) |
| Antibiotic treatment — no. (%) | 4(33.3%) | 3(30.0%) |
| Antifungal treatment — no. (%) | 0 | 0 |
| Antiviral treatment — no. (%) | 12(100.0%) | 10(100.0%) |
| Glucocorticoids — no. (%) | 12(100.0%) | 10(100.0%) |
| Intravenous Ig therapy — no.(%) | 2(16.7%) | 2(20.0%) |
| **Clinical outcome** |  |  |
| Remained in hospital — no. (%) | 4 (33.3%) | 7 (70.0%) |
| Discharged — no. (%) | 7 (58.4%) | 2 (20.0%) |
| Discharged for severe cases— no. (%) | 3 (50.0%) |  |
| Discharged for mild cases— no. (%) | 4 (100%) |  |
| Died — no. (%) | 1 (8.3%) | 1 (10.0%) |

Two weeks after initiation of DIP treatment, 3 of the 6 severe cases (50%) and all 4 of the mild cases (100%) were discharged from the hospital. One of the critically ill patient with extremely high levels of D-dimer and lymphopenia at the time of receiving DIP passed away. All other patients were in clinical remission. In comparison, the discharge and remission rates of the mild and severe cases were inferior in the control patients (**Table 2**).

**Table 2.** Clinical outcomes of 27 enrolled patients.

| Table 2. Clinical outcomes of 27 enrolled patients |  |  |  |
| --- | --- | --- | --- |
|  | Severity of illness<br>— no. (%) | Outcomes<br>(up to 2/26) | Total discharge<br>— no. (%) |
| Dipyridamole group<br>(n=12) | Mild — 4 (33.3%) | 4 discharged (100%) | 7 (58.4%) |
|  | Severe — 6 (50.0%) | 3 discharged (50%)<br>2 in remission (33%) |  |
|  | Critical ill — 4 (33.3%) | 1 in remission (50%)<br>1 death (2/09) |  |
| Control group (n=10) | Mild — 4 (40.0%) | 3 discharged (75%) | 4 (40.0%) |
|  | Severe — 4 (40.0%) | 1 discharged (25%)<br>1 in remission (25%) |  |
|  | Critical ill — 2 (20.0%) | 1 death (2/18) |  |

The baseline temperature was higher and the oxygenation status was worse in the DIP-treated group than those in the control group, however, temperature and oxygenation were stabilized in the mild and severe patients from both groups after one day of treatment (**Figure S2**). Furthermore, we observed a continuously increased trend in lymphocyte counts and significantly increased platelet numbers in patients receiving DIP treatment in comparison to the control group (**Figure 3A**). Given that lymphocytopenia and thrombocytopenia are common in the severe and critically ill patients, immune recovery may contribute to infection resolution in DIP-treated patients. It should be noted that 4 patients from the DIP-treatment group and 2 patients from the control group had increased baseline levels of D-dimer when they were admitted to the hospital (**Table 1**). We measured dynamic changes in reference to the respective baseline levels for all patients, and found that the levels of D-dimer continuously rose in the control group, whereas they were decreased and stabilized in the DIP-treated group (**Figure 3A, Figure S2**).

**Figure 3.**
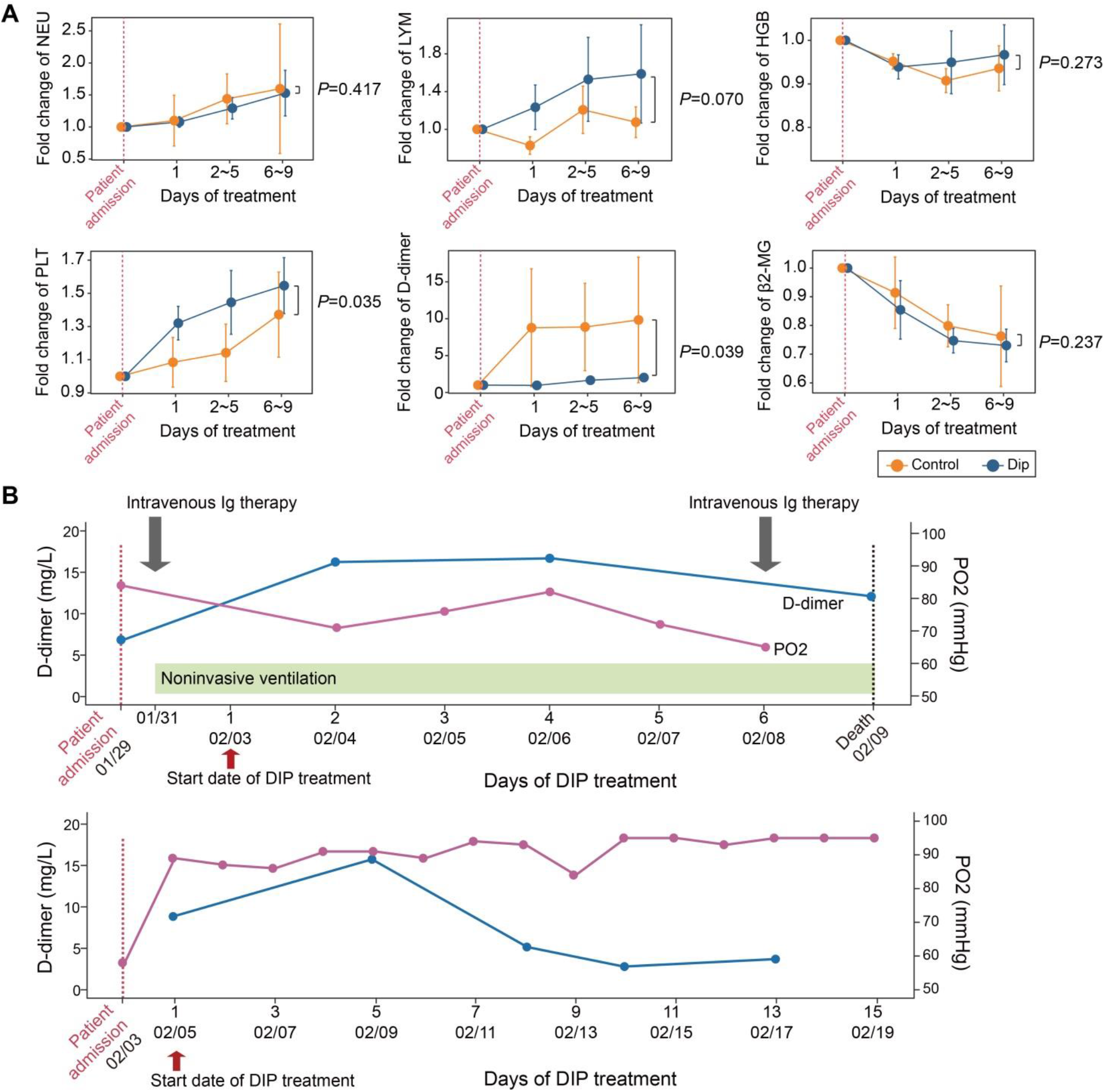
Changes of the study variables during the course of treatment. (A) Dynamic changes in the routine blood indexes (NEU, LYM, HGB, and PLT), coagulation variable (D-dimer), and kidney function indices (b2-MG) in reference to the baseline values. The levels of D-dimer were sustained in DIP treatment group, while those in the control group were significantly increased. Lymphocyte counts were persistently elevated in trend and PLT counts was significantly increased in the DIP group in comparison to the control group. Data are shown as the means ± SE across different time bins during the treatment course. *P* values were calculated by Student’s *t* test and indicated beside each panel. (B) Schematics of the treatment overview and clinical parameters of the deceased critically ill patient (top) and the surviving patient (bottom) who received DIP adjunctive therapy.

We then examined the two critically ill patients who received DIP adjunctive therapy. A 70-year-old man with very low oxygen saturation and has suffered from hypoxia and multiorgan dysfunction at hospital administration has passed away. He had extremely high levels of D-dimer (16.2 mg/L) and very low lymphocyte count (0.37×10^9^/L) at the time of receiving DIP adjunctive therapy. His oxygen saturation remained low and has unfortunately passed away 5 days since initiation of DIP treatment. In contrast, the other severely ill patient also had very low oxygen saturation and high D-dimer level (8.83 mg/L) at administration. His D-dimer level had gone up to as high as 15.72 mg/L, but gradually declined to 3.69 mg/L following DIP adjunctive therapy. The patient has since been in clinical remission. This reinforces that high levels of D-dimer and low lymphocyte counts are associated with poor prognosis and suggest that that DIP treatment should be initiated before the progression to critical illness (**Figure 3B**).

All patients had chest CT scans and showed typical multiple patchy ground-glass shadows in the lungs before the treatment. In the DIP treated patients, the lesions had varied degree of absorption. Typical findings following DIP treatment was illustrated in one of the severe COVID-19 patient who had taken sequential CT scans (**Figure 4)**.

**Figure 4.**
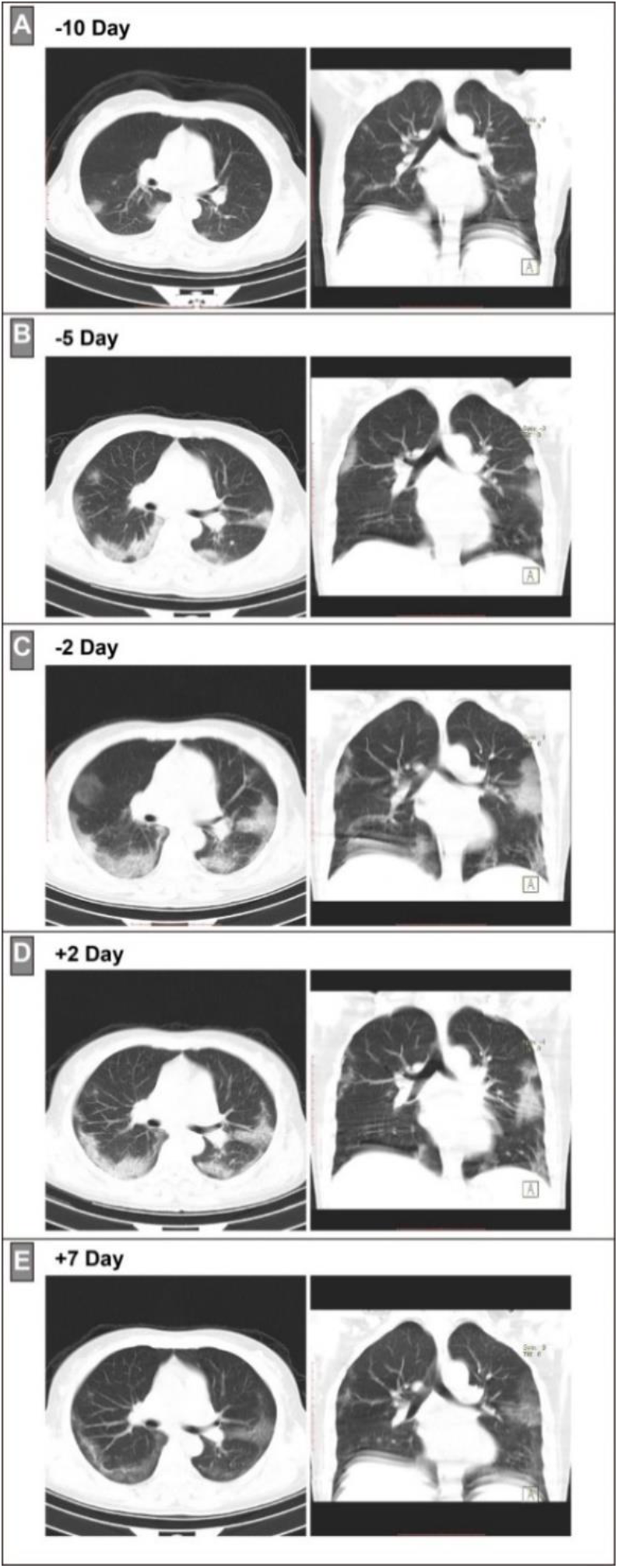
Chest CT images in the axial (left panel) and coronal view (right panel) of represented severe COVID-19 patient before and after receiving DIP adjunctive therapy. (A) Initial chest CT scan 10 days before DIP treatment. (B) Chest CT scan 5 days before DIP treatment. (C) Chest CT scan 2 days before DIP treatment. (D) Chest CT scan 2 days after DIP treatment. (E) Chest CT scan 7 days after DIP treatment.

## Discussion

Despite the enormous threat of HCoV-19, no drugs have been claimed to be effective including the existing drugs used to treat other viruses. In reference to SARS-CoV-infection, we hypothesized that the HCoV-19 Spike protein engagement may activate RAS in the lung.^6,43^ This hypothesis was supported by published clinical characteristics and biochemical indices of the severe and critically ill COVID-19 patients, who showed ARDS, hypertension, acute heart, kidney injury, and positive D-dimer results.^2,19,20^ In searching for available anticoagulants, we focused on DIP because of its broad-spectrum antiviral, anti-inflammatory, and anti-fibrotic effects. We confirmed that DIP possessed potent antiviral immunity to single-stranded RNA viruses *in vitro*, and in a VSV-induced viral pneumonia model *in vivo*. Most importantly, a laboratory confirmed EC50 of 0.1 mM to suppress HCoV-19 replication highly suggested that the therapeutic dosage of DIP may potentiate effective antiviral responses in infected patients. These findings are in concordance with the overall remarkable outcomes in patients receiving one week of DIP adjunctive therapy, seven of the twelve patients were discharged from the hospital and four achieved clinical remission.

We found that DIP adjunctive treatment was effective in preventing hypercoagulability if applied early in the severe COVID-19 patients. DIP treatment blunted the increase in D-dimer levels and increased circulating platelet and leucocytes counts, and thus was associated with overall remarkable clinical cure and remission rates. High levels of D-dimer closely correlated with pulmonary embolism,^44^ vascular thrombosis, and renal dysfunction.^45^ It is an important prognostic factor of whether ICU-patients may recover from severe infections.^46,47^ As such, the antiviral, anti-inflammatory, and anti-hypercoagulability effects of DIP could reduce the risk of micro-vascular thrombosis in HCoV-19 infected patients. This could in turn improve vascular circulation and restore vital functions of important organs such as heart and kidney.

In summary, we suggest anti-coagulant therapy for COVID-19 patients with early signs of elevated D-dimer levels. DIP is an attractive anti-coagulant adjunctive therapy due to its safe clinical profile as an anti-platelet agent. It also holds additional benefits as a pan-PDE inhibitor in eliciting broad-spectrum antiviral immunity, as well as anti-inflammatory and anti-fibrotic effects. Moreover, the wide availability, safety and affordability of DIP argue for further investigation into its therapeutic use in COVID-19, particularly in the event of its rapid spread into the developing counties.

## Data Availability

The authors confirm that the data supporting the findings of this study are available within the article [and/or] its supplementary materials.

## Contributors

JZ, XH, YZ, FZ, and HBL co-designed the study and co-led overall data interpretation. ZL led the virtual screening, YH led the SPR analysis, ZC, ZZ, JW, YX, JL, HX, RF, FB, CJ, HH, and CO also participated in study design and performed *in vivo* and *in vitro* studies. XL, SL, YS, BX, XH, and FZ led the clinical analysis. XL, SL, YS, BX, XH, and FZ participated in data collection. ZC participated in data analysis. XL, SL, and ZC produced the tables and figures. AML reviewed the manuscript. YZ and HBL drafted the manuscript with significant input from JZ, XH, and FZ. All authors interpreted the results and critically revised the manuscript for scientific content. All authors approved the final version of the Article.

## Acknowledgments

We cordially acknowledge Tencent Cloud, National Supercomputing centers in Guangzhou and Shenzhen, and SenseTime for providing HPC resources for virtual screening and free energy perturbation calculations. We cordially thank Prof. H. Ke at the University of North Carolina, Chapel Hill, for his help to improve our writing of this work.

